# The High Price of Equity in Pulse Oximetry: A cost evaluation and need for interim solutions

**DOI:** 10.1101/2023.09.21.23295939

**Authors:** Katelyn Dempsey, Mary Lindsay, James E. Tcheng, An-Kwok Ian Wong

**Author notes:** **Corresponding Author An-Kwok Ian Wong**, *Assistant Professor of Medicine, Department of Medicine, Division of Pulmonary, Allergy, and Critical Care Medicine, Department of Biostatistics and Bioinformatics, Division of Translational Biomedical Informatics*, 2 Genome Ct, Box 103000, Duke University, MSRB 2, Durham, NC 27710.

## Abstract

**Importance:** Disparities in pulse oximetry accuracy, disproportionately affecting patients of color, have been associated with serious clinical outcomes. Although many have called for pulse oximetry hardware replacement, the cost associated with this replacement is not known.

**Objective:** To estimate the cost of replacing all pulse oximetry hardware throughout a hospital system.

**Design:** Single-center survey, 2023

**Setting:** Single center.

**Participants:** One academic medical center with three hospitals.

**Main Outcomes and Measures:** Cost of fleet replacement as identified by current day prices for hardware.

**Results:** New and used prices for 5,079/5,678 (89.5%) across three hospitals for pulse oximetry devices were found. The average equipment cost to replace pulse oximetry hardware is $15,704.12 per bed. Replacement and integration costs are estimated at $28.5-31.8 million for the entire medical system. Extrapolating these costs to 5,564 hospitals in the United States results in an estimated cost of $14.1 billion.

**Conclusions and Relevance:** “Simply replacing” pulse oximetry hardware to address disparities may be neither simple, cheap, or timely. Solutions for addressing pulse oximetry accuracy disparities leveraging current technology may be necessary.

**Trial Registration:** Pro00113724, exempt

**Key points:** *Question:* What is the cost and complexity of replacing pulse oximetry technology to improve disparities, both at a single institution and across the US?

*Findings:* In this observational study of pulse oximetry devices in an academic medical system with three hospitals, new and used prices were found for 5,079/5,678 devices (89.5%), with fleet replacement and integration cost of $28.5-31.8 million and some life cycles extending beyond 18 years. When extrapolated to 5,564 hospitals in the United States, estimated replacement costs are $9.7-$20.1 billion.

*Meaning:* The monetary and time cost of pulse oximetry hardware replacement is substantial, and solutions utilizing current pulse oximetry technology are essential to delivering equitable care to all patients.

## Background

Pulse oximetry is a critical tool in modern medicine.^1^ Patients of color - especially Black patients - have been associated with increased pulse oximetry accuracy disparities, as evidenced by increased hidden or occult hypoxemia.^2,3^ These inaccuracies are associated with increased mortality, organ dysfunction, decreased oxygen therapy, and delayed COVID recognition.^3–5^ Many have called for simply replacing all pulse oximetry devices with new technology. There are early possibilities, such as green light oximetry, cameras, and improved algorithms.^6–8^ However, “simple replacement” may neither be simple, cheap, nor timely. The objective of this manuscript is to describe the financial and logistical burden in replacing pulse oximeters at one health system and across the United States. ^2,6^

### Types of pulse oximetry equipment

Pulse oximetry devices fall into three categories:

- **Multi-Parameter Models** (e.g., Philips MX800, GE Dash 4000): Complex units that display multiple vital signs, predominantly used in ICUs and operating rooms.
- **Pulse Ox Modules/Monitors** (e.g., Masimo Rad-7): Specialized for pulse oximetry, these can be integrated into existing systems without replacing whole units.
- **Vital Signs Monitors** (e.g., Philips SureSigns VS4): Primarily used in outpatient and general inpatient settings, these collect various vital signs.

Monitors usually interface with electronic medical record systems. Standalone units require manual data entry. Among these, pulse ox modules are the easiest to replace.

## Methods

### Equipment data

The Hospital System X provided extensive equipment inventories, categorized by individual locations. New and used equipment prices were sought online, primarily utilizing Google and comprehensive sites like Bimedis.com. Price data was retrieved in June 2023. All prices were calculated in United States dollars. Data on hospital size was retrieved from the American Hospital Directory. To reconcile new and used prices, the lowest ratio of new or Manufacturer’s Suggested Retail Prices (MSRP) to used prices was conservatively estimated from devices with both new and used prices. No discount rate was assumed given the lack of transparency to discount rates.

The Duke University Health System IRB approved Protocol Pro00113724 as exempt.

### Device integration estimates

The integration process involves multiple variables, including server specifications, firewalls, and software revisions. The time required was estimated at 250-500 person-hours per device model, at a rate of $200/hour. Costs were fully applied to multi-parameter monitors, 50% to pulse ox modules/monitors, and not applied to vital signs monitors.

### Extrapolation to United States National Hospital data

Hospital data from 2015 was obtained from the Health, United States data finder.^10^ Fleet replacement costs were modeled in two ways: based on estimated costs per bed by hospital A-C and based on used price percentages of MSRP.

We utilized 2015 national hospital data from the Health, United States data finder. Estimates for fleet replacement costs were based on two models: one using cost per bed by types of hospitals and the other using used price percentages of MSRP.

## Results

### Cost estimates for a single academic health system

Hospital System X comprises three main facilities. Hospital A is a large center with 1,048 beds, including 332 designated for specialized care. Hospital B is a medium-sized facility with 186 beds, of which 28 are specialized. Hospital C has 373 beds, with 22 allocated for specialized care. Across these hospitals, 140 distinct types of pulse oximetry equipment are in use. Available pricing data cover 85.9% of all 8,460 devices in the system, and 67.1% of these are actively in use within the hospitals.

Two key devices, Philips SureSigns VS4 and MX700, have both new and used pricing available. The ratio of new-to-used prices for these devices ranges from 513% to 133%. Consequently, the new Manufacturer’s Suggested Retail Price (MSRP) was conservatively approximated as 133% of the used prices, with an adjustment range of 110% to 200%. Of the 5,678 devices currently in hospital use, 89.5% had either new or used price data. The estimated replacement cost for these devices, not including integration costs, is $25.2 million. This estimate encompasses 89.5% of devices currently in use, with per-bed costs ranging from $10,801 to $22,392.

Additional costs for integrating these devices are projected to be between $3.25 and $6.5 million, leading to a total projected cost of $28.5 to $31.8 million for the hospitals. When extending the scope to include clinics affiliated with Hospital System X, the estimated replacement cost for all devices rises to $35.9 million. With projected integration costs of an additional $4.4 to $8.8 million, the overall expenditure is estimated to be between $40.3 and $44.7 million.

### Extrapolation to national costs

National United States hospital data from the CDC 2015 data reveals 5,564 hospitals, with an average of 161.4 beds/hospital. Estimates looking at different base hospital rates, assuming 75%, are in Table 2. At an estimated cost from $10,801-22,392/bed for fleet replacement, these estimates suggest that fleet replacement will cost $9.7-20.1 billion.

**Table 1.** Average replacement cost for all beds in a hospital across the system. Based on the number of devices with prices, this table reflects the cost of replacing pulse oximeters by individual hospitals, along with across the three hospitals together. Prices for replacement cost are in thousands (e.g., Hospital A $17.1 million, B $4.2 million, C $4.0 million).

|  |  | A | B | C | Health System X |
| --- | --- | --- | --- | --- | --- |
| total number of devices |  | 3769 | 919 | 990 | 5678 |
| devices with prices |  | 3,339 (88.6%) | 829 (90.2%) | 911 (92.%) | 5,079 (89.5%) |
| number of beds |  | 1,048 | 186 | 373 | 1,607 |
| special care beds |  | 332 | 28 | 22 | 382 |
| operating rooms |  | 53 | 18 | 20 | 91 |
| number of models | Multi-Parameter Module/Monitor | 39 | 24 | 13 | 48 |
|  | Pulse Ox Module/Monitor | 28 | 9 | 5 | 34 |
|  | Vital Signs Monitor | 53 | 9 | 8 | 58 |
| replacement cost (thousands) | | \$ 17,055.58 | \$ 4,164.96 | \$ 4,028.88 | \$ 25,236.52 |
| average replacement cost per bed | | \$ 16,274.41 | \$ 22,392.28 | \$ 10,801.30 | \$ 15,704.12 |

**Table 2.**
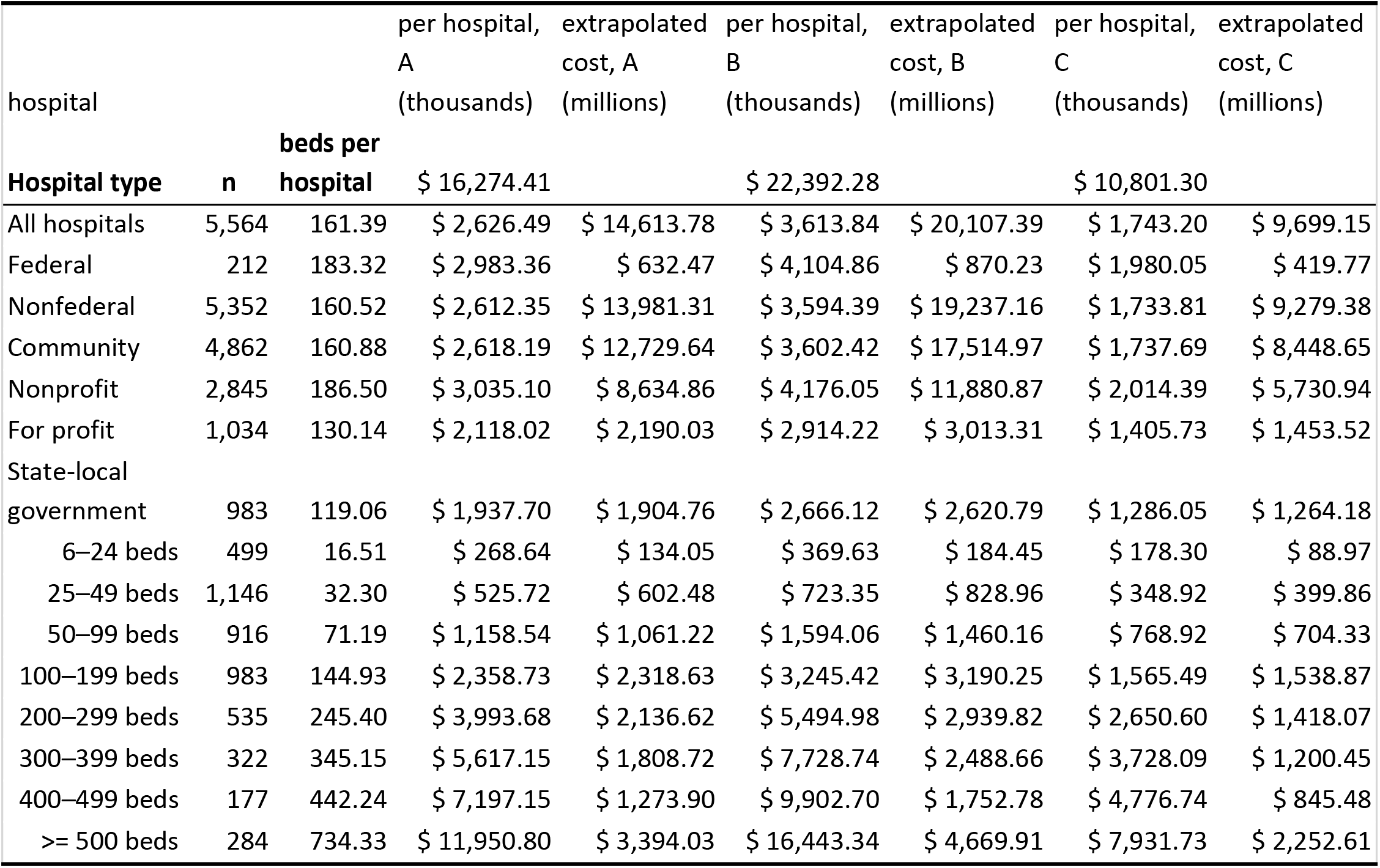
National estimates in fleet replacement cost, using different hospital types for estimates, using 75% used/new price estimates. This table reflects the extrapolated cost of replacing pulse oximetry equipment for all 5,564 hospitals in the 2015 CDC hospital data chart. This model explores a range of costs for replacing pulse oximetry per bed from a quaternary academic medical center (hospital A) through community hospitals (B, C). This does not include integration costs.

Using an average of all rates across the health system, the new MSRP can be estimated from used costs. Based on an estimated cost of $15,704/bed ($13,727-22,085/bed), the estimated fleet replacement cost is $14.1 billion ($13.7-22.1 billion). (Supplemental Table 4)

## Discussion

The racial disparities in pulse oximetry accuracy are not just statistically alarming but also clinically urgent. Such disparities contribute to increased morbidity and mortality among patients of color, a pressing issue that cannot be relegated to the lengthy timelines associated with natural equipment replacement cycles.^9^ Our analysis reveals that full equipment overhaul is neither cost-effective nor timely, thereby mandating prioritized attention at federal and industry levels for alternative solutions.

There is an immediate need for targeted funding and policies that can drive rapid innovation in this space. Federal agencies must take the lead in establishing frameworks that incentivize the development of more accurate, yet economically viable, pulse oximetry technologies. Similarly, industry stakeholders should prioritize research and development investments towards creating lower-cost solutions that can be deployed quickly.

Informatics-based approaches are especially promising in this context. By leveraging existing hardware and data analytics, we can implement more accurate and less invasive solutions sooner. These strategies not only avoid the massive financial burden of full-scale equipment replacement—estimated up to $22.1 billion nationally—but also present an opportunity for quicker implementation. However, such approaches would benefit substantially from federal and industry support, be it in the form of research grants, tax incentives, or regulatory fast-tracking.

### Limitations

The study has limitations, such as incomplete price data and unaccounted volume discounts, which may underestimate the actual replacement costs. Also, device integration costs may be underestimated, particularly for vital signs monitors.

## Conclusion

The racial disparities in pulse oximetry accuracy have immediate and dire clinical ramifications. A simplistic notion of device replacement is neither affordable nor timely, demanding an estimated $9.7-22.1 billion and several years for nationwide implementation. Given these challenges, it is essential to prioritize federal and industry interventions that can innovate lower-cost and quicker solutions, such as informatics-based approaches. Policymakers, healthcare providers, and researchers must urgently collaborate to navigate this complex problem efficiently and equitably.

## Data Availability

We can make percentages available for units above 1% prevalence.

---

**Figure 1.**
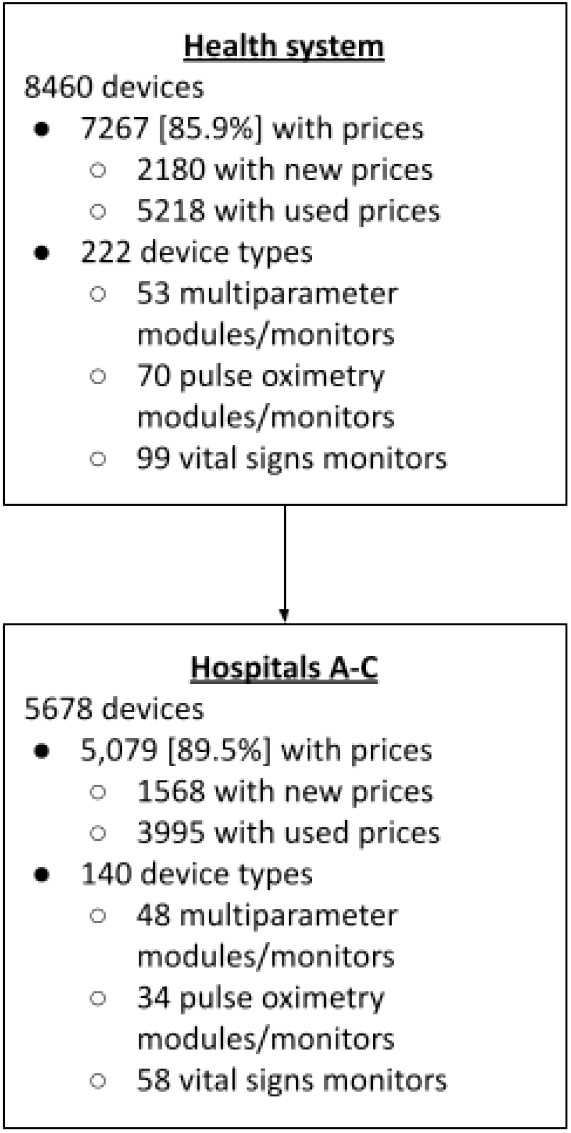
Flow diagram.

## References

1. Priority medical devices list for the COVID-19 response and associated technical specifications [Internet]. 2020 [cited 2023 Jul 31];Available from: https://www.who.int/publications/i/item/WHO-2019-nCoV-MedDev-TS-O2T.V2

2. Sjoding MW, Dickson RP, Iwashyna TJ, Gay SE, Valley TS. Racial Bias in Pulse Oximetry Measurement. N Engl J Med 2020;383(25):2477–8.

3. Wong AI, Charpignon M, Kim H, et al. Analysis of discrepancies between pulse oximetry and arterial oxygen saturation measurements by Race/Ethnicity and association with organ dysfunction and mortality. JAMA Network Open [Internet] 2021;Available from: 10.1001/jamanetworkopen.2021.31674

4. Henry NR, Hanson AC, Schulte PJ, et al. Disparities in Hypoxemia Detection by Pulse Oximetry Across Self-Identified Racial Groups and Associations With Clinical Outcomes* [Internet]. Critical Care Medicine. 2022;50(2):204–11. Available from: 10.1097/ccm.0000000000005394

5. Gottlieb ER, Ziegler J, Morley K, Rush B, Celi LA. Assessment of Racial and Ethnic Differences in Oxygen Supplementation Among Patients in the Intensive Care Unit. JAMA Intern Med [Internet] 2022 [cited 2022 Jul 11];Available from: https://jamanetwork.com/journals/jamainternalmedicine/article-abstract/2794196

6. Raposo A, Silva R, Rosário LB, Sanches J, da Silva HP. Smartphone Pulse Oximetry Using Two-tone Camera-based Photoplethysmography [Internet]. [cited 2023 Aug 8];Available from: https://ieeexplore.ieee.org/abstract/document/10175332

7. Gokhale SG, Daggubati V, Alexandrakis G. Innovative technology to eliminate the racial bias in non-invasive, point-of-care (POC) haemoglobin and pulse oximetry measurements. BMJ Innovations [Internet] 2023 [cited 2023 Aug 8];9(2). Available from: https://innovations.bmj.com/content/9/2/73.abstract

8. Berwal D, Kuruba A, Shaikh AM, Udupa A, Baghini MS. SpO2 Measurement: Non-Idealities and Ways to Improve Estimation Accuracy in Wearable Pulse Oximeters [Internet]. [cited 2023 Aug 8];Available from: https://ieeexplore.ieee.org/abstract/document/9762716

9. Seo G, Park S, Lee M. How to calculate the life cycle of high-risk medical devices for patient safety. Front Public Health 2022;10:989320.

10. Data finder - Health, United States [Internet]. 2023 [cited 2023 Jul 11];Available from: https://www.cdc.gov/nchs/hus/data-finder.htm?year=2017&table=Table%20089

11. Rowling SC, Fløjstrup M, Henriksen DP, et al. Arterial blood gas analysis: as safe as we think? A multicentre historical cohort study. ERJ Open Res [Internet] 2022;8(1). Available from: 10.1183/23120541.00535-2021

